# A brief review of antiviral drugs evaluated in registered clinical trials for COVID-19

**DOI:** 10.1101/2020.03.18.20038190

**Authors:** Drifa Belhadi, Nathan Peiffer-Smadja, François-Xavier Lescure, Yazdan Yazdanpanah, France Mentré, Cédric Laouénan

**Affiliations:** Université de Paris, IAME, INSERM, F-75018 Paris, France; Département d’Epidémiologie Biostatistiques et Recherche Clinique, Hôpital Bichat – Claude-Bernard, AP-HP, 75018 Paris, France; Infectious Diseases Department, Bichat-Claude Bernard Hospital, Assistance-Publique Hôpitaux de Paris, Paris, France

**Keywords:** COVID-19, SARS-CoV-2, coronavirus, clinical trials, review

## Abstract

**Background:** Although a number of antiviral agents have been evaluated for coronaviruses there are no approved drugs available. To provide an overview of the landscape of therapeutic research for COVID-19, we conducted a review of registered clinical trials.

**Methods:** A review of currently registered clinical trials was performed on registries, including the Chinese (chictr.org.cn) and US (clinicaltrials.gov) databases to identify relevant studies up to March, 7^th^ 2020. The search was conducted using the search terms “2019-nCoV”, “COVID-19”, “SARS-CoV-2”, “Hcov-19”, “new coronavirus”, “novel coronavirus”. We included interventional clinical trials focusing on patients with COVID-19 and assessing antiviral drugs or agents.

**Findings:** Out of the 353 studies identified, 115 clinical trials were selected for data extraction. Phase IV trials were the most commonly reported study type (n=27, 23%). However, 62 trials (54%) did not describe the phase of the study. Eighty percent (n=92) of the trials were randomized with parallel assignment and the median number of planned inclusions was 63 (IQR, 36-120). Open-label studies were the most frequent (46%) followed by double-blind (13%) and single blind studies (10%). The most frequently assessed therapies were: stem cells therapy (n=23 trials), lopinavir/ritonavir (n=15), chloroquine (n=11), umifenovir (n=9), hydroxychloroquine (n=7), plasma treatment (n=7), favipiravir (n=7), methylprednisolone (n=5), and remdesivir (n=5). Remdesivir was tested in 5 trials with a median of 400 (IQR, 394-453) planned inclusions per trial, while stem cells therapy was tested in 23 trials, but had a median of 40 (IQR, 23-60) planned inclusions per trial. Lopinavir/ritonavir was associated with the highest total number of planned inclusions (2606) followed by remdesivir (2155). Only 52% of the clinical trials reported the treatment dose (n=60) and only 34% (n=39) the duration. The primary outcome was clinical in 76 studies (66%), virological in 27 (23%); radiological in 9 (8%) or immunological in three studies (3%).

**Interpretation:** Numerous clinical trials have been registered since the beginning of the COVID-19 outbreak, however, a number of information regarding drugs or trial design were lacking.

**Funding:** None

## MANUSCRIPT

### INTRODUCTION

On December 31^st^, 2019, the first suspected cases of an epidemic of viral pneumonia of unknown aetiology were reported in the city of Wuhan, China. Patients were linked to Huanan market, selling fish and other live animals. On January 7^th^, 2020 the Chinese health authorities and the World Health Organization (WHO) officially announced the discovery of a novel coronavirus, currently called SARS-CoV-2.^1^ The disease caused by SARS-CoV-2 has been named COVID-19. On January 30^th^, WHO declared the epidemic a Public Health Emergency of International Concern (PHEIC). On March 11^th^, WHO characterized COVID-19 as a pandemic. Up to March the 18^th^, more than 200,000 cases of COVID-19 and more than 8,000 deaths have been reported in the world. The COVID-19 epidemic is unique because of its scale, the speed of its spread, the lack of pre-existing scientific data and the importance of media and scientific coverage.^2^ The scientific and public health community have responded with early publication of clinical data and predictions of spread and guidance for effective containment.^3,4^ Alongside, and critically, health professionals need to find effective and safe treatment for patients infected with SARS-CoV-2.

### Why is this review needed?

Although a number of drugs have been evaluated for SARS-CoV and MERS-CoV there are no approved therapeutic agents available for coronaviruses. Integrating clinical trials of experimental therapeutics is an increasingly recognized part of the response during infectious disease outbreaks. Since the Ebola outbreak in West Africa and subsequent outbreaks in the Democratic Republic of Congo, clinical trials of investigative drugs have been fully integrated in the epidemic response.^5–7^ The COVID-19 pandemic is unique because of its scale, the speed of its spread, the lack of pre-existing scientific data and the importance of media and scientific coverage.^2^ To encourage the development of clinical trials that test therapeutics against SARS-CoV-2, the WHO has suggested a number of candidate antiviral agents that have to be tested in clinical trials.^8^ Concomitantly, numerous clinical trials have been registered since the beginning of the COVID-19 outbreak numerous to evaluate therapeutic strategies for this disease.

In the epidemic context, it is crucial for clinicians and researchers to have access to rapid and quality information on clinical trials that the various teams around the world are setting up. The results will inform about the antiviral agents that are used, their dosing and duration, the inclusion and exclusion criteria for patients, the outcomes that will be evaluated, as well as the design of the clinical trials.

We believe that it is essential to carry out a review of these early phase clinical trials before the results are even available in order to best inform the teams wishing to test new therapies, choose therapeutic candidates or to design clinical trials.

In this review, we aimed to summarize the current state of registered clinical trials for COVID-19 in order to study their design, which antiviral agents were being investigated, the details of their administration, and the outcomes.

## METHODS

### Search strategy and selection criteria

A search was performed on clinical trial registries of privately and publicly funded clinical trials worldwide. We selected the following clinical trial registries: U.S. (https://clinicaltrials.gov/), Chinese (www.chictr.org.cn/), Korean (https://cris.nih.go.kr/cris/en/), Iranian (https://www.irct.ir/), Japanese (https://rctportal.niph.go.jp/en/), and European (https://www.clinicaltrialsregister.eu/). We chose these locations as they were the ones with the highest number of cases at the time of extraction. We added the WHO clinical trial registry (http://apps.who.int/trialsearch/) and the International Standard Randomised Controlled Trial Number (ISRCTN) Registry, recognized by the WHO and the International Committee of Medical Journal Editors (ICMJE). A first search was conducted on February 28^th^, 2020 on all registries listed above to capture the studies registered from November 2019; and an updated search was conducted on March 7^th^, 2020 on clinicaltrials.gov to capture additional studies registered since February 28^th^. Our search strategy was designed to identify all the clinical trials using antiviral agents that were registered for COVID-19. The following search terms were used for our search to capture relevant studies: “2019-nCoV”, “COVID-19”, “SARS-CoV-2”, “Hcov-19”, “new coronavirus”, “novel coronavirus”. Data extracted from selected studies included study design, sponsorship, population, outcomes, and inclusion/exclusion criteria.

The eligibility criteria were developed using the Patient Intervention Comparison Outcomes Study type (PICOS) framework^9^.

Inclusion criteria were:

- Population: patients with COVID-19,
- Intervention/Comparator: any antiviral agent or drug. We excluded trials evaluating traditional Chinese medicine, homeopathy, dietary supplements, and therapeutic strategies whose description was not sufficient to identify a specific drug.
- Outcomes: any outcomes,
- Study type: interventional clinical trial.

We excluded traditional Chinese medicine and homeopathy as we have no expertise to analyse clinical trials testing these agents that rely on controversial concepts.^10,11^ Dietary supplements were also excluded as their potential in treating COVID-19 seems limited.

## Definitions

We considered clinical trials between those planning to include only patients with severe diseases, those planning to include patients with moderate diseases and those planning to include both. We defined severe patients as patients requiring either non-invasive ventilation, high flow oxygen devices or invasive mechanical ventilation or extracorporeal membrane oxygenation (ECMO). Patients with moderate pneumonia were patients who did not require these. Studies were further analysed according to the primary endpoint, that could be clinical, virological (viral excretion in clinical samples), radiological (imaging results such as CT-scan or X-rays), or immunological (CD8+/CD4+ T cells count, IFN-gamma measurement results).

## RESULTS

### Number of studies

Our search identified 353 studies, and 115 clinical trials were selected for data extraction in the review (Figure 1). Among the 238 excluded studies, 125 were trials that did not focus on an intervention of interest, e.g. traditional Chinese medicine, and 81 were not interventional clinical trials. Among the 115 included clinical trials, 39 were registered on the US clinical trial registry and 76 on the Chinese clinical trial registry. Sponsorship was not systematically reported in the Chinese clinical trial registry preventing us from accurately analysing private or public sponsorship. We found one trial with planned inclusions in the USA (NCT04280705) while the others recruited patients in China but data were lacking in many trials.

**Figure 1:**
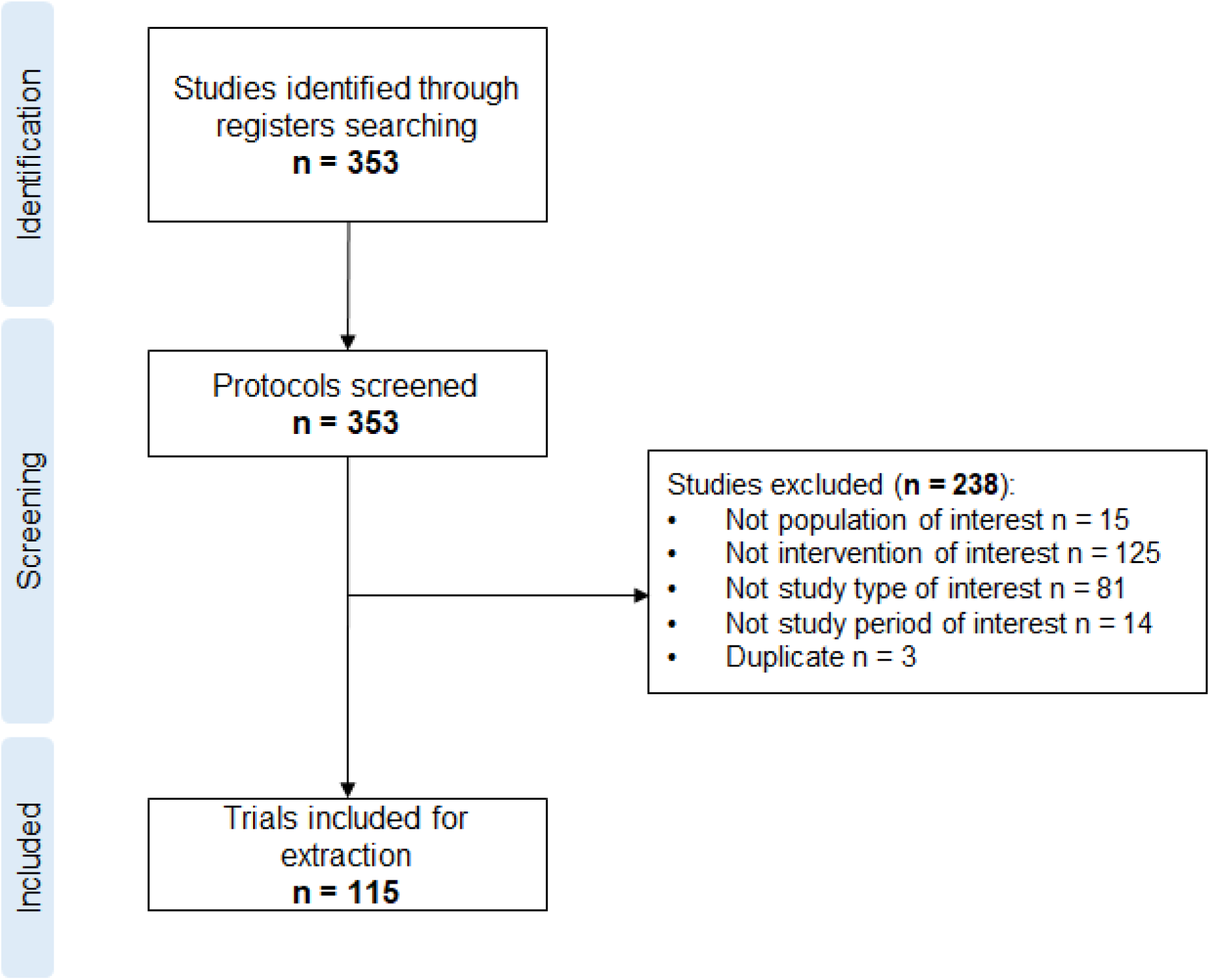
Selection process of clinical trials.

### Population and severity of disease

Children were included in two clinical trials in China, one testing darunavir with cobicistat (without age precision, NCT04252274) and one on human menstrual blood-derived stem cells (1 to 99 years old, ChiCTR2000029606). Six other planned to recruit patients aged over 15: one testing lopinavir/ritonavir and favipiravir plus alpha-Interferon atomization (ChiCTR2000029600); one hydroxycholoroquine (ChiCTR2000029740); one convalescent plasma treatment (ChiCTR2000029850); one recombinant human granulocyte-colony stimulating factor (G-CSF) (ChiCTR2000030007); one favipiravir (ChiCTR2000030113), and one human mesenchymal stem cells (ChiCTR2000030138). All the registered trials excluded pregnant women. The trials were evenly divided between the patients with moderate pneumonia (n=29, 25%), moderate or severe pneumonia (n=57, 50%), and severe pneumonia (n=29, 25%; Table 1).

**Table 1:** Description of the clinical trials registered for the treatment of COVID-19

|  |  | N = 115 (%) |
| --- | --- | --- |
| Study phase |  |  |
|  | Phase I | 4 (3) |
|  | Phase II* | 13 (14) |
|  | Phase III** | 9 (8) |
|  | Phase IV | 27 (23) |
|  | Unspecified | 62 (54) |
| Disease severity |  |  |
|  | Moderate infection only | 29 (25) |
|  | Severe or moderate infection | 57 (50) |
|  | Severe infection only | 29 (25) |
| Study design |  |  |
|  | Randomized | 92 (80) |
|  | Non-randomized | 12 (11) |
|  | Single-arm | 11 (10) |
| Blinding |  |  |
|  | Double-blind | 15 (13) |
|  | Single-blind | 11 (10) |
|  | Open-label | 53 (46) |
|  | Non-applicable† | 6 (5) |
|  | Unspecified | 30 (26) |
| Total number of planned inclusions |  |  |
|  | <50 | 40 (35) |
|  | 50-100 | 28 (24) |
|  | 100-150 | 21 (18) |
|  | 150-200 | 3 (3) |
|  | 200-250 | 7 (6) |
|  | ≥250 | 16 (14) |
| Primary endpoint |  |  |
|  | Clinical | 76 (66) |
|  | Virological | 27 (23) |
|  | Radiological | 9 (8) |
|  | Immunological | 3 (3) |

\*Including phase I/II trials;
\*\*Including phase II/III trials;
†: Single-arm or factorial trials.

### Studies design

Phase IV trials were the most commonly reported study type (n=27, 23%; Table 1) before phase II (n=13, 14%) and phase III (n=9, 8%). However, most of the registered trials did not describe the phase of the study (n=62, 54%). Regarding blinding, we found 53 open-label studies (46%), 15 double-blind (13%), and 11 single-blind (10%). The vast majority of trials were randomized (n=92, 80%) with a parallel assignment between arms. The median (IQR) number of planned inclusions was 63 (36-120) with a range of 9 to 600 participants.

### Treatments

Various treatments were evaluated in the clinical trials, the most frequently evaluated ones are described in Table 2. Only 52% of the clinical trials reported the treatment dose (n=60) and only 34% (n=39) the duration. A table with the detailed combination therapies and the identification of each clinical trial is available in the Supplementary material. Figure 2 reports the number of trials by the median of the total number of planned inclusions per trial for the ten most frequent therapies (stem cells therapy, lopinavir/ritonavir, chloroquine phosphate, hydroxychloroquine, favipiravir, umifenovir, plasma treatment, remdesivir, methylprednisolone, oseltamivir). Remdesivir was tested in only 5 trials, but these trials had the highest median number of planned inclusions per trial (400, IQR 394-453). At the other end of the spectrum, stem cells therapy was associated with the highest number of trials (23 trials), but with a small median number of planned inclusions per trial (40, IQR 23-60). Figure 3 shows the total number of planned inclusions and the number of clinical trials for the ten most frequently assessed treatments. Lopinavir/ritonavir was associated with the highest total number (2606) followed by remdesivir (2155) and umifenovir (1705).

**Table 2:** Description of the antiviral agents evaluated in more than one clinical trial registered for COVID-19

| Treatment | Number of randomised trials/Number of trials* | Dose** | Duration** | Median number of planned inclusions (IQR) | Severity of the disease† |
| --- | --- | --- | --- | --- | --- |
| Stem Cells therapy | 18/23 | NA | 5-7 days | 40 (23-60) | Moderate or severe |
| Lopinavir/Ritonavir | 11/15 | 400mg/100mg oral twice a day | 7-14 days | 120 (80-183) | Moderate or severe |
| Chloroquine phosphate | 8/11 | 500mg oral twice a day | 10 days | 100 (90-116) | Moderate or severe |
| Umifenovir | 7/7 | 200mg oral 3 times a day | 14 days | 125 (80-390) | Moderate or severe |
| Hydroxychloroquine | 7/7 | 200mg oral twice a day | 14 days | 100 (89-220) | Moderate or severe |
| Plasma treatment | 3/7 | 200-500ml IV | NA | 95 (38-100) | Moderate or severe |
| Favipiravir | 6/7 | 1600-2400mg oral loading dosage then 200-600mg oral twice a day | 10-14 days | 60 (30-75) | Moderate or severe |
| Methylprednisolone | 5/5 | 1mg/kg/day IV | 3-7 days | 100 (80-100) | Moderate or severe |
| Remdesivir | 5/5 | 200mg IV loading dose on day 1, followed by 100mg IV once-daily | 5-10 days | 400 (394-453) | Moderate or severe |
| Oseltamivir | 2/2 | 75mg oral once or twice a day | 14 days | 230 (145-315) | Moderate or severe |
| Baloxavir Marboxil | 2/2 | 80mg oral on day 1, on day 4, and day 7 | 7 days | 30 (30-30) | Moderate |
| Thalidomide | 2/2 | 100 mg oral per day | 14 days | 70 (40-100) | Moderate or severe |
| Darunavir/cobicistat | 2/2 | 800mg/150mg oral per day | NA | 65 (30-100) | Moderate or severe |
| Thymosin | 2/2 | 1.6mg SC once a day | 5 days | 120 (120-120) | Severe |
| PD-1 blocking antibody | 2/2 | 200mg IV, one time | 1 time | 80 (40-120) | Severe |
| Tocilizumab | 1/2 | NA | NA | 124 (60-188) | Moderate or severe |
| Intravenous Immunoglobulin | 1/2 | 200-500mg/kg/d IV | 3-5 days | 45 (10-80) | Severe |
| Ozonated autohemotherapy | 1/2 | NA | NA | 60 (60-60) | Moderate or severe |
| Type 1 Interferon injection | 1/1 | NA | NA | 30 (-) | Moderate or severe |
| Interferon nebulization | 1/1 | NA | NA | 100 (-) | Moderate or severe |

\*: We added trials using the drug alone or as part of a combination therapy
\*\*: The most frequent among trials was selected
†: We defined severe patients as patients requiring either non-invasive ventilation, high flow oxygen devices or invasive mechanical ventilation or extracorporeal membrane oxygenation (ECMO). Patients with moderate pneumonia were patients who did not require these.
Note: IV: intravenous; SC: subcutaneous;

**Figure 2:**
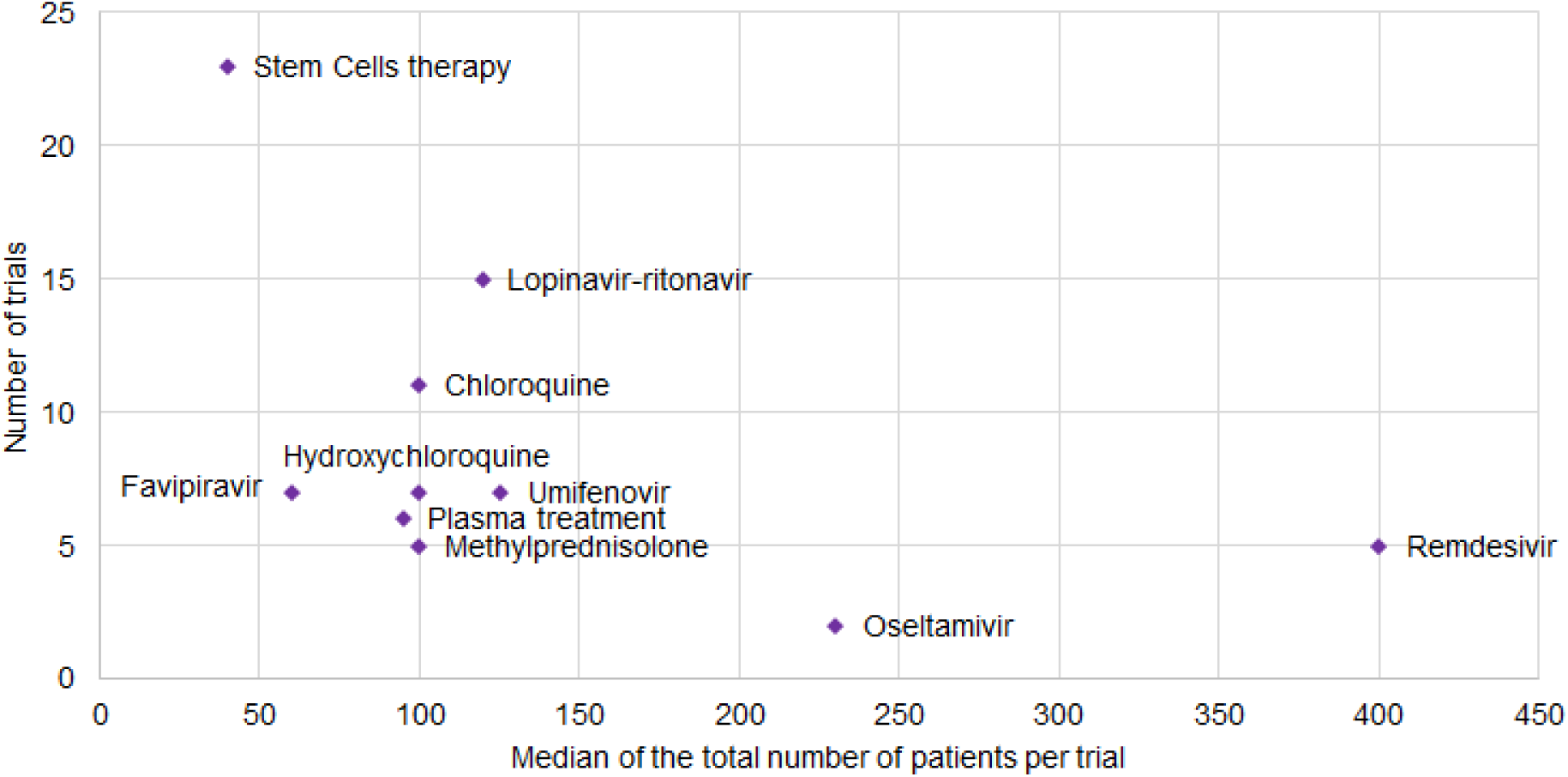
Number of trials reported by the median of the total number of planned inclusions per trial for the most common treatments.

**Figure 3:**
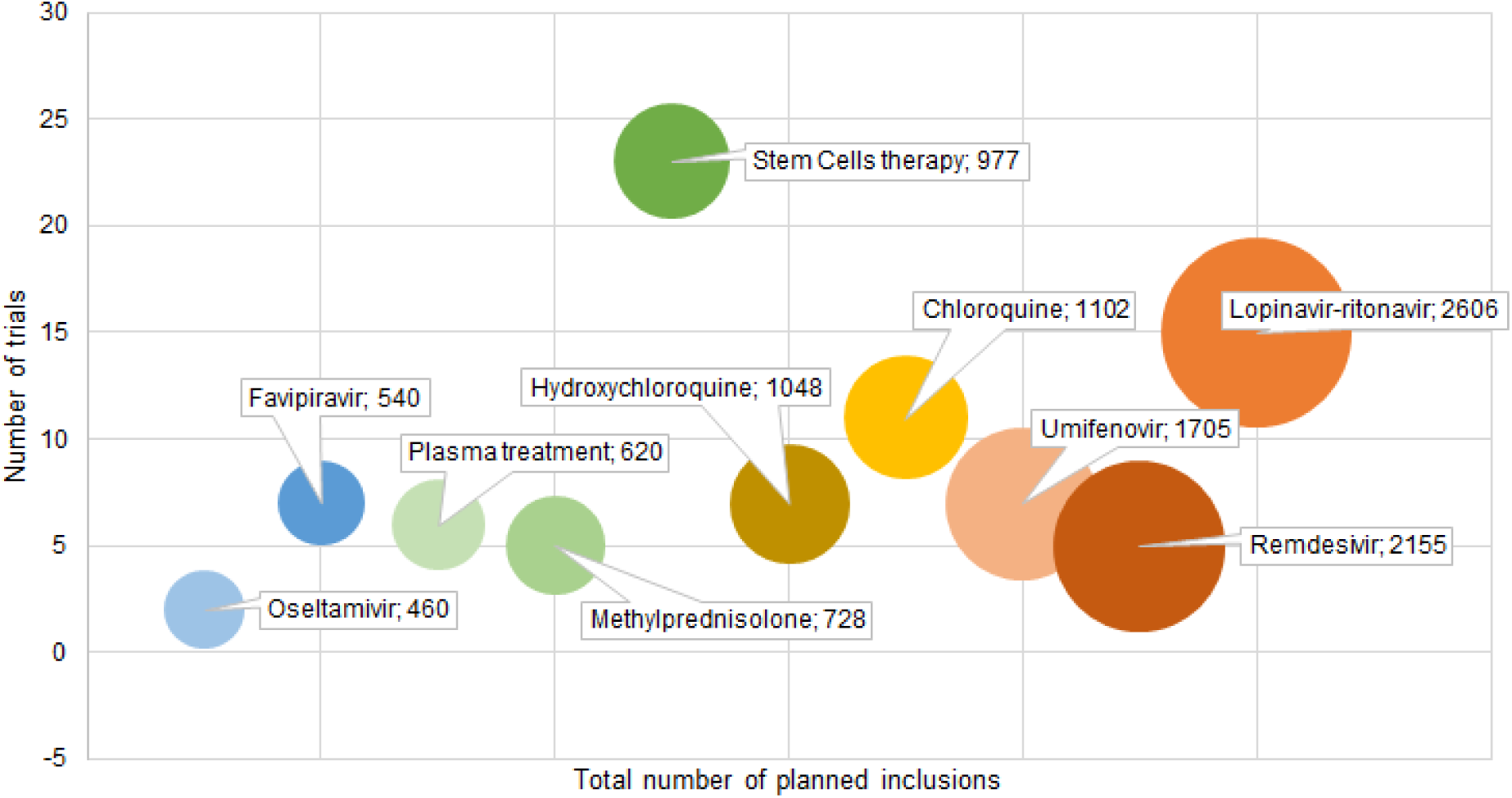
Number of trials per total number of planned inclusions (in all the trials) for the ten most frequently assessed treatments. The size of the circle corresponds to the addition of the total numbers of planned inclusions for all trials evaluating one of the treatments

### Endpoints

The primary outcome was clinical in 76 studies (66%; Table 1), most of them focused on the evolution of the symptoms such as the time to clinical recovery, the proportion of patients with clinical improvement or deterioration, the length of hospitalization or the mortality. A number of scores were used as a primary outcome such as the ordinal 7-point scale adapted from WHO master protocol^8^ (10 studies), the lung injury score^12^ (4 studies), the pneumonia severity index^13^ (3 studies) or the National Early Warning Score (NEWS) 2 score (2 studies).^14^ In other studies, the primary outcome was either virological in 27 studies (23%), radiological in nine studies (8%) or immunological in three studies (3%).

## DISCUSSION

Based on the evidence available up to March 7^th^, 2020, stem cells therapy and lopinavir-ritonavir were the most frequently evaluated candidate therapies in terms of number of trials (23 and 15 trials respectively), whereas remdesivir was associated with the highest median number of planned inclusions per trial (400, IQR 394-453)) for 5 trials only.

This review of ongoing clinical trials assessing COVID-19 treatments shows the important amount of research that is currently being conducted on this topic. However, although the number of trials identified is high, there are a number of caveats.

First, numerous treatments have been selected based on various levels of supporting preclinical data. Most of the agents evaluated in clinical trials have shown an *in vitro* antiviral activity, sometimes including coronaviruses. Lopinavir/ritonavir is tested in 15 clinical trials in this review. This combination has shown an *in vitro* activity against SARS-CoV in several studies^15^ and appears to have activity against MERS-CoV in animal studies^16^. The use of this agent for treatment of COVID-19 has been described in case reports^17,18^ and in a case series of patients infected with SARS-CoV-2 in Singapore.^19^ We found 18 clinical trials evaluating hydroxychloroquine or chloroquine, whose mechanism of action is similar.^20^ The *in vitro* antiviral activity of chloroquine has been known for a long time^21^ and was described on a number of viruses including SARS-CoV.^22^ However, chloroquine failed to demonstrate a benefit in the treatment of viral diseases such as influenza, dengue or chikungunya.^23–25^ Regarding COVID-19, a recent publication reported an activity of chloroquine on SARS-CoV-2^26^ and another encouraged the use of chloroquine for patients with COVID-19 on the basis of unreported clinical results.^27^ Experts in China have suggested the use of chloroquine for patients infected with SARS-CoV-2 but no clinical data has been provided yet to support this announcement.^28,29^ Remdesivir is evaluated in 5 clinical trials but with the highest median number of planned inclusions per trial. Studies *in vitro* in human airway epithelial cell assays demonstrated that remdesivir inhibits replication of coronaviruses, including MERS-CoV.^30^ In mouse infection models, remdesivir had therapeutic efficacy against SARS-CoV and MERS-CoV.^30,31^ In a recent non-human primate study, remdesivir treatment initiated 12 hours post inoculation with MERS-CoV provided clinical benefit with a reduction in clinical signs, reduced virus replication in the lungs, and decreased presence and severity of lung lesions.^32,33^ The rationale of using stem cell therapy is based on its immunomodulatory properties that could be interesting in severe COVID-19.^34,35^ However, stem cell-based therapies have not demonstrated an effect in treating other viral diseases and the scientific background to test them is weak. The fact that stem cell therapy, that has not shown any effect in antiviral diseases, was one of the most frequently assessed therapies is unexpected. This highlights the fact that researchers should strive to conduct clinical trials with the most promising candidates, according to *in vitro* and preclinical *in vivo* scientific data.

Second, data is often lacking regarding study designs and on the treatment being assessed, such as the dose and duration. This restricts the information available for researchers and potentially delay the finding of successful treatments. Third, most of the trials planned including a low number of patients, which reduces the robustness of the future results of those clinical trials. Although, these numbers should be taken with caution as they only represent an anticipated number of inclusions for each trial and not the actual number of patients included. Fourth, primary outcomes were very heterogeneous in the clinical trials. The use of clinical outcomes should be encouraged in a disease for which we do not know the association between viral clearance, radiological or immunological evolution and clinical status.

Due to the pandemic context associated with COVID-19, the number of clinical trials registered is increasing day after day. A previous review conducted up to February 18^th^, 2020 found 74 clinical trials evaluating antiviral agents or drugs.^36^ Our review adds to this evidence by screening a larger number of clinical trial registries and reporting the studies design, randomization, allocation, and number of planned inclusions as well as treatment dose, duration, disease severity, and primary outcomes used.

This important amount of work conducted by researchers is encouraging for the therapeutic research for this new disease. However, care should be taken when designing a clinical trial in this complicated context as robust results are needed in order to be able to find the appropriate treatment. Finding the best agents for rapid implementation in clinical trials for a new outbreak is challenging. Our study underlines the need to register as much details as possible on clinical trials registries during outbreaks in order to inform the development of future trials. The scientific background supporting the use of a treatment should be clear and detailed as much as possible. The dose and duration of drugs evaluated, as well as details on the study design, the population of interest and the primary outcome, are crucial elements that have to be shared in the context of the epidemic response. Reporting as much details as possible is key to have consistent clinical trials and to enhance the reproducibility of the results, especially as studies are more often associated with a low number of planned inclusions and composite or weak outcomes that can limit the efficacy assessment of the treatments. That is why transparency and consistency are crucial when reporting clinical trials in order to improve statistical power by conducting, for example, meta-analyses.

The development of clinical trials during an outbreak is an adaptive process and new evidence is produced at an impressive rate. A review of the strategies that are already registered in official clinical registries of clinical trials is an important asset for researchers and methodologists. These results might inform the adaptation of existing clinical trials and the development of additional trials.

## Data Availability

The authors confirm that the data supporting the findings of this study are available within the article and its supplementary materials.

## AUTHORS CONTRIBUTIONS

Conceptualization: Belhadi D, Peiffer-Smadja N, Yazdanpanah Y, Mentre F, Laouénan C Conduct of the review: Belhadi D, Peiffer-Smadja N

Writing original draft: Belhadi D, Peiffer-Smadja N

Writing - review and editing: Belhadi D, Peiffer-Smadja N, Yazdanpanah Y, Mentre F, Laouénan C

## CONFLICTS OF INTEREST

YY is the chair of the Global Research Collaboration for Infectious Disease Preparedness (GloPID-R) and the coordinator of REsearch and ACTion targeting emerging infectious diseases (REACTing). We declare no competing interests.

## Notes

### Competing Interest Statement

The authors have declared no competing interest.

### Funding Statement

No funding.

